# Prediction of elective surgical red blood cell transfusion reveals surgeon-level variation

**DOI:** 10.64898/2026.11.03.26364952

**Authors:** Esa Turkulainen, Mikko Arvas, Miika Koskinen, Minna Ilmakunnas

## Abstract

**BACKGROUND:** Preoperative prediction of red blood cell (RBC) transfusion in elective surgery serves two complementary purposes: enabling patient blood management (PBM) interventions before surgery and supporting blood product supply planning for elective lists. Beyond patient risk factors, individual surgeon practice may also contribute to transfusion rates, but this effect has not been fully quantified.

**STUDY DESIGN AND METHODS:** In a retrospective cohort of 279,905 elective surgical procedures at Helsinki University Hospital (2021–2025), we trained four gradient-boosted tree (CatBoost) models crossing patient factors with procedure group and pseudonymised surgeon identity. All predictors were restricted to data available seven or more days before scheduled surgery. The primary outcome was any allogeneic RBC transfusion from operating-room entry through seven postoperative days. Discrimination was assessed by the area under the precision-recall curve (AUPRC) on a held-out 20% test set. Surgeon-level effects were characterised by Shapley additive explanation (SHAP) values.

**RESULTS:** The transfusion rate was 2.2%. The patient-only model achieved AUPRC 0.261 (no-skill baseline 0.022). Adding surgeon identity to the procedure-adjusted model increased AUPRC from 0.440 to 0.472 (+0.032; 95% CI 0.020–0.044), raising lift from 20.3 to 21.7. Surgeon identity was the third most important of 72 features by permutation importance.

**DISCUSSION:** The operating surgeon was associated with RBC transfusion beyond patient and procedure factors, and surgeon identity was the third most important predictor in a deployment-realistic model using only data available a week before surgery. This information is readily available from routine records and could support PBM audit and blood supply planning for elective surgery.

## INTRODUCTION

Perioperative red blood cell (RBC) transfusion is among the most common interventions in elective surgery. Although transfusion is frequently necessary, the current AABB guidelines recommend restrictive policies, because transfusion carries well-characterised hazards including acute and delayed transfusion reactions, transmission of infection, and transfusion-related immunomodulation.^1^ In surgical patients, allogeneic transfusion is also independently associated with adverse postoperative outcomes, including wound infection, major cardiac and thromboembolic events, and reduced survival.^2–5^ Restrictive transfusion policies are supported by patient blood management (PBM) programmes, which allow for correction of anaemia, optimization of anticoagulation, and haemostatic preparation prior to surgery. However, PBM relies on the identification of patients with high risk of perioperative bleeding and transfusion.

In addition to improving patient outcomes, anticipating blood use in advance supports blood supply planning, whereby blood banks and transfusion services allocate inventory in advance of scheduled surgical lists. Traditionally, elective blood use is anticipated in hospitals coarsely based on the procedure type and its historical risk of bleeding, and a handful of patient variables, such as preoperative haemoglobin (Hb) levels. With the increasing use of electronic health records (EHR), these estimates can now draw on more granular patient and procedure information, reducing wastage and operational costs.

Preoperative prediction of RBC transfusion has been attempted for more than three decades, but performance has plateaued. A systematic review and meta-analysis of prognostic models for perioperative RBC transfusion in adult cardiac surgery identified nine externally validated models developed since 1989 and 27 external validation studies. Only two models, TRUST and TRACK, had been externally validated at least five times. Their summary c-statistics were 0.74 (95% CI 0.65–0.81) and 0.72 (0.68–0.75).^6^ A parallel review spanning elective surgery of all types reached the same conclusion, reporting c-statistics of 0.67–0.78 across 72 developed and 48 externally validated models.^7^

None of these models incorporated clinician- or hospital-level factors.^6,8^ Successive generations have instead recombined the same patient-level predictors: preoperative haemoglobin or red cell mass, body size, sex, age, renal function, and a marker of anticipated procedural difficulty, and have converged on a similar performance ceiling.^8,9^ In a two-centre cohort of 816,618 operations, a 24-predictor preoperative model built from this familiar set reached a c-statistic of 0.93 but a positive predictive value of only 8.9% at a 2.3% transfusion rate, and was non-inferior to three machine-learning–derived scores.^10^ This ceiling thus appears to reflect the limits of patient characteristics rather than of the modelling approach. The same review that documented this plateau recommended incorporating physician- and hospital-level information as a route forward.^6^

Yet substantial evidence across surgical specialties indicates that the operating clinician is itself a determinant of transfusion. After adjustment for patient characteristics and procedure type, transfusion rates vary significantly between surgeons and between hospitals.^11–22^ The differences are attributable to intraoperative haemostatic techniques, clinicians’ tolerance of intraoperative blood loss, and postoperative transfusion-trigger thresholds rather than to case-mix.^22^ Because surgeon identity is known when surgery is scheduled, this variation could make transfusion predictions for individual procedures more accurate, supporting both PBM and blood supply planning for elective lists.

Testing this requires a model that adjusts for both patient case mix and procedure complexity. Gradient-boosted decision trees such as CatBoost^23^ suit this task: they handle high-cardinality categorical variables such as surgeon identity via a special ordered target encoding.

In this study, we trained four CatBoost models in a two-by-two factorial design to test whether surgeon identity improves the prediction of elective surgical RBC transfusion beyond patient and procedure factors. We used a seven-day predictive pre-operative horizon to ensure that the predictions are operationally useful in PBM and supply chain management settings.

## MATERIALS AND METHODS

### Study design and setting

This was a retrospective cohort study at Helsinki University Hospital (HUS), which is responsible for specialist care for the Uusimaa region of Finland (approximately 1.7 million inhabitants) through a network of hospitals, and is also a national tertiary referral centre. The cohort therefore covers almost all elective specialist surgery in the region. The study included all elective surgical procedures performed during January 2021 – December 2025. The research and data permit for this study was granted by the HUS institutional review board (HUS/579/2022, 17 October 2022). Pursuant to the Act on the Secondary Use of Health and Social Data (26 April 2019/552) in Finland, individual patient consent was waived given the registry-based design. Reporting follows the TRIPOD+AI guidelines (Supplementary Checklist).

### Data sources and study population

Patient and procedural data were extracted from the hospital EHR (Apotti, Epic Systems Corporation) system by HUS data lake personnel and delivered to a secure analysis environment HUS Acamedic.

Only elective procedures, identified by national urgency codes, in adult (≥18 years) patients were included, with each procedure treated as the unit of analysis. Procedures were excluded if operating-room entry time, exit time, or primary procedure code (Nordic Classification of Surgical Procedures, NCSP) was missing. Procedures were grouped by the first three characters of their primary NCSP code (the procedure group), which identify the anatomical region and type of procedure; for example, JJB comprises liver resections. To obtain stable procedure-level estimates, we excluded primary procedure codes occurring fewer than 50 times during the study period. Patients were excluded if they had missing or non-binary sex, or if their ASA physical status was class V (indicating that either the ASA class or the elective status of the procedure had been coded erroneously). Procedures with a missing ASA class were retained, with missing class treated as a separate category.

The operating clinician of each procedure was recorded as a pseudonymised identifier. Where several clinicians were recorded for a procedure, a primary surgeon was selected by seniority of professional title, with ties broken at random. In this paper, we refer to the operating clinician as ‘surgeon’ regardless of the operative specialty. No minimum number of procedures per surgeon was required.

### Predictive horizon and look-back windows

All predictors were derived exclusively from data recorded at least seven days before the scheduled operating-room entry (the predictive horizon), the point at which PBM interventions and blood supply planning remain feasible. Look-back windows preceding the horizon were domain-specific: twelve months for comorbidity diagnoses; 10, 30 or 90 days for medications depending on drug class (Supplementary Table S1); and unbounded for laboratory results and prior bleeding events. For laboratory results, the time elapsed since the qualifying measurement was included as a predictor instead of an age cut-off (see Predictors).

### Outcome

The prediction target for each elective procedure was the transfusion of at least one unit of allogeneic RBCs. A transfusion was attributed to the procedure if it was started between operating-room entry and seven days after operating-room exit. The window was closed earlier at hospital discharge or at the patient’s next operation, elective or emergency, so that each transfusion was attributed to at most one procedure.

### Predictors

From patient demographics we captured sex and age at surgery. From patient measurements we collected body mass index (calculated from height and weight) and estimated blood volume (using formulas for non-obese and obese adults).^24,25^ From procedure data we collected the ASA physical status class (I–IV, or missing).

For preoperative laboratory values, we extracted the most recent result recorded before the predictive horizon for each of the following: haemoglobin (g/L), platelet count (×10⁹/L), creatinine (µmol/L), C-reactive protein (mg/L), RBC mean corpuscular volume (MCV; K), international normalised ratio (INR), albumin (g/L), leucocyte count (×10⁹/L), and ferritin (µg/L), and, for each, the number of days between the measurement and operating-room entry. Where no direct INR measurement was available, INR was derived from the thromboplastin time percentage (TT%) through a log–log regression fitted on paired same-draw measurements (Supplementary Methods, section 3 and Supplementary Figure S1). 78.7% of the 127,273 INR values in the cohort were derived in this way. Values were left missing where no qualifying result existed.

Comorbidities were identified from ICD-10-coded hospital visits within the twelve-month look-back window, using the 31 Elixhauser categories of the Quan et al. coding algorithm ^26^ (Supplementary Table S2). Each category was encoded as the number of visits on which it was recorded. In addition, an overall comorbidity burden score (the van Walraven-weighted Elixhauser index^27^) was calculated per visit, and its maximum within the window was included as a single numeric predictor.

Medication exposure was encoded as binary indicators for twelve drug classes, derived from Anatomical Therapeutic Chemical coded (ATC) prescriptions in the EHR systems. A class was flagged if any prescription in it was active during its class-specific look-back window before the horizon (drug classes, ATC codes and windows in Supplementary Table S1). The number of prescription records active in the current EHR system within 30 days of the horizon was included as a proxy for polypharmacy.

Prior bleeding was summarised as the cumulative recorded blood loss from all earlier surgical episodes before the horizon, including intraoperative (weighed swabs, suction canister volumes) and postoperative (drain output, estimated dressing loss) bleeding.

### Data preprocessing

Preoperative laboratory values were not imputed, and no missingness-indicator variables were added. Whether a test was ordered is itself informative in EHR data,^28,29^ and CatBoost can use this without a substitute value.^30^ Missing body mass index and blood volume (7.2%) were regression-imputed (Supplementary Methods, section 2).

### Model development

Four CatBoost classification models were trained in a two-by-two factorial design. They shared the same patient features and differed only in whether procedure group and surgeon identity were added as categorical predictors: the patient-only model (patient features only), the patient + procedure model (adding procedure group), the patient + surgeon model (adding surgeon identity) and the full model (patient + procedure + surgeon). Each model outputs a predicted probability of transfusion for every procedure, and all four were evaluated by the area under the precision-recall curve (AUPRC) on the same held-out test set. Because the models differ only in the grouping variables, the difference in AUPRC between two models measures the predictive information added by the variable that distinguishes them. The patient + procedure and patient + surgeon models, compared with the patient-only model, give the unadjusted contributions of procedure and surgeon. The full model, compared with the patient+ procedure model, gives the contribution of surgeon identity beyond procedure, which was the primary comparison.

Patients were randomly assigned to a training set or a held-out test set, with all of a patient’s procedures kept in the same set to prevent leakage of patient-level information into the test set. The split was stratified by whether the patient had any transfused procedure and targeted 20% of procedures in the test set (Supplementary Figure S2). Hyperparameters (learning rate, tree depth, L2 leaf regularisation) were selected within the training set by a 30-draw random search^31^ with five-fold cross-validation, with folds grouped by patient, optimising AUPRC. Class weighting was not applied: in an ablation within the training set, inverse-frequency weighting did not improve AUPRC and produced substantially worse-calibrated probabilities (Supplementary Table S3).

### Statistical analysis

Because transfusion was uncommon, AUPRC was the primary performance metric.^32^ Unlike the area under the receiver operating characteristic curve, it focuses on how well the rare transfused cases are identified, and its no-skill value equals the outcome prevalence rather than 0.5. The primary comparison was the gain in AUPRC from adding surgeon identity to the procedure-adjusted model. We also report lift (AUPRC divided by the no-skill baseline) and the share of all transfusions among the 5% of test-set procedures with the highest predicted risk. 95% confidence intervals for all these measures were obtained by paired cluster bootstrap resampling patients in the test set (2,000 resamples).

Feature importance in the full model was assessed by permutation feature importance on the test set, defined as the decrease in AUPRC when a feature’s values are randomly shuffled, averaged over 20 permutations. As a sensitivity analysis, surgeon identity was also shuffled only within procedure groups.

SHAP values^33^ were used to describe surgeon-specific effects. SHAP values decompose each individual prediction into additive per-feature contributions. A positive contribution for a given feature indicates that the feature raised the model’s predicted risk of transfusion for that procedure above the average prediction. Contributions are expressed on the log-odds scale. The surgeon effect was defined as each surgeon’s mean SHAP contribution for surgeon identity across their test-set procedures, among surgeons with at least 15 test-set procedures. Surgeons whose mean SHAP contribution was more than 1 SD (of the distribution of surgeon effects) above or below zero were described as having above- or below-average surgeon effects. Surgeon effects were checked against observed test-set transfusions (Supplementary Methods, section 10). In exploratory analyses among the same surgeons, we related the surgeon effect to surgeon volume (total number of procedures in the study period; Spearman correlation with a bootstrap 95% CI) and to seniority of professional title.

Calibration was assessed by Brier score, calibration intercept and slope, and for the full model by reliability diagram. Full-model performance was also assessed by sex and age. All analyses were performed in R^34^ version 4.6.1. fley packages were data.table^35^ 1.18.4 (data manipulation), catboost^23^ 1.2.10 (model training and SHAP computation), and PRROC^36^ 1.4 (precision-recall curve estimation). Code is published openly on GitHub, and an accompanying snapshot is archived on Zenodo.^37^

## RESULTS

### Cohort characteristics

The analysed cohort comprised 279,905 elective surgical procedures in 205,985 unique patients, performed by 1,543 surgeons across 294 procedure groups (Table 1; Supplementary Figure S2). The median (IQR) number of procedures per surgeon was 69 (9–242), and 27 surgeons, with one test-set procedure each, appeared only in the test set. The overall transfusion rate was 2.2% (6,133 procedures; 95% CI 2.1–2.2%). The median (IQR) preoperative haemoglobin was 136 (126–146) g/L. 31.1% of procedures had no qualifying preoperative haemoglobin result.

**Table 1.**

| Variable | All | Transfused | Not transfused |
|---|---|---|---|
| N procedures | 279,905 | 6,133 | 273,772 |
| Age, years, median (IQR) | 64 (48-75) | 70 (59-77) | 64 (48-75) |
| Sex, Male | 124,320 (44.4%) | 2,490 (40.6%) | 121,830 (44.5%) |
| Sex, Female | 155,585 (55.6%) | 3,643 (59.4%) | 151,942 (55.5%) |
| ASA class, 1 | 35,697 (12.8%) | 167 (2.7%) | 35,530 (13.0%) |
| ASA class, 2 | 126,731 (45.3%) | 1,592 (26.0%) | 125,139 (45.7%) |
| ASA class, 3 | 84,919 (30.3%) | 2,939 (47.9%) | 81,980 (29.9%) |
| ASA class, 4 | 11,345 (4.1%) | 1,420 (23.2%) | 9,925 (3.6%) |
| ASA class, Missing | 21,213 (7.6%) | 15 (0.2%) | 21,198 (7.7%) |
| BMI, median (IQR) | 27 (24-30) | 26 (23-29) | 27 (24-30) |
| Estimated blood volume, L, median (IQR) | 4.8 (4.1-5.5) | 4.6 (4.0-5.3) | 4.9 (4.1-5.5) |
| Preoperative Hb, g/L, median (IQR) | 136 (126-146) | 119 (105-132) | 137 (126-146) |
| Platelet count, ×10<sup>9</sup>/L, median (IQR) | 250 (208-298) | 264 (209-332) | 250 (208-297) |
| No qualifying preoperative Hb, n (%) | 87,147 (31.1%) | 655 (10.7%) | 86,492 (31.6%) |
| Transfusion rate, n (%, 95% CI) | 6,133 (2.2%, 2.1-2.2%) |  |  |
| Top 5 transfused procedure groups, n (%) |  |  |  |
| Primary prosthetic replacement of hip joint (NFB) | 6,240 (2.2%) | 587 (9.6%) | 5,653 (2.1%) |
| Excision of rectum (JGB) | 2,117 (0.8%) | 371 (6.0%) | 1,746 (0.6%) |
| Total excision of uterus (LCD) | 5,834 (2.1%) | 357 (5.8%) | 5,477 (2.0%) |
| Primary prosthetic replacement of knee joint (NGB) | 8,702 (3.1%) | 344 (5.6%) | 8,358 (3.1%) |
| Connection to coronary artery from internal mammary artery (FNA) | 835 (0.3%) | 289 (4.7%) | 546 (0.2%) |

### Model discrimination

A model without predictive ability would have an AUPRC equal to the test-set transfusion rate (no-skill baseline 0.022), and all four models exceeded this substantially (Table 2, Figure 1). The patient-only model achieved AUPRC 0.261, 12 times the baseline. Adding procedure group increased AUPRC to 0.440 (+0.178; 95% CI 0.156–0.201), and adding surgeon identity alone increased it to 0.384 (+0.123; 95% CI 0.103–0.144). The full model achieved AUPRC 0.472 (+0.210; 95% CI 0.185–0.235). After adjustment for procedure group, adding surgeon identity increased AUPRC by 0.032 (95% CI 0.020–0.044; from 0.440 to 0.472), corresponding to an increase in lift from 20.3 to 21.7 (7% relative gain; 95% CI 4–10%). If the 5% of procedures with the highest predicted risk were flagged, the full model would identify 68.9% (95% CI 66.2– 71.4%) of all transfusions, compared with 66.6% (63.9–69.3%) for the patient + procedure model (10 additional transfused cases per 1,000 flagged; 95% CI 2–17).

**Figure 1.**
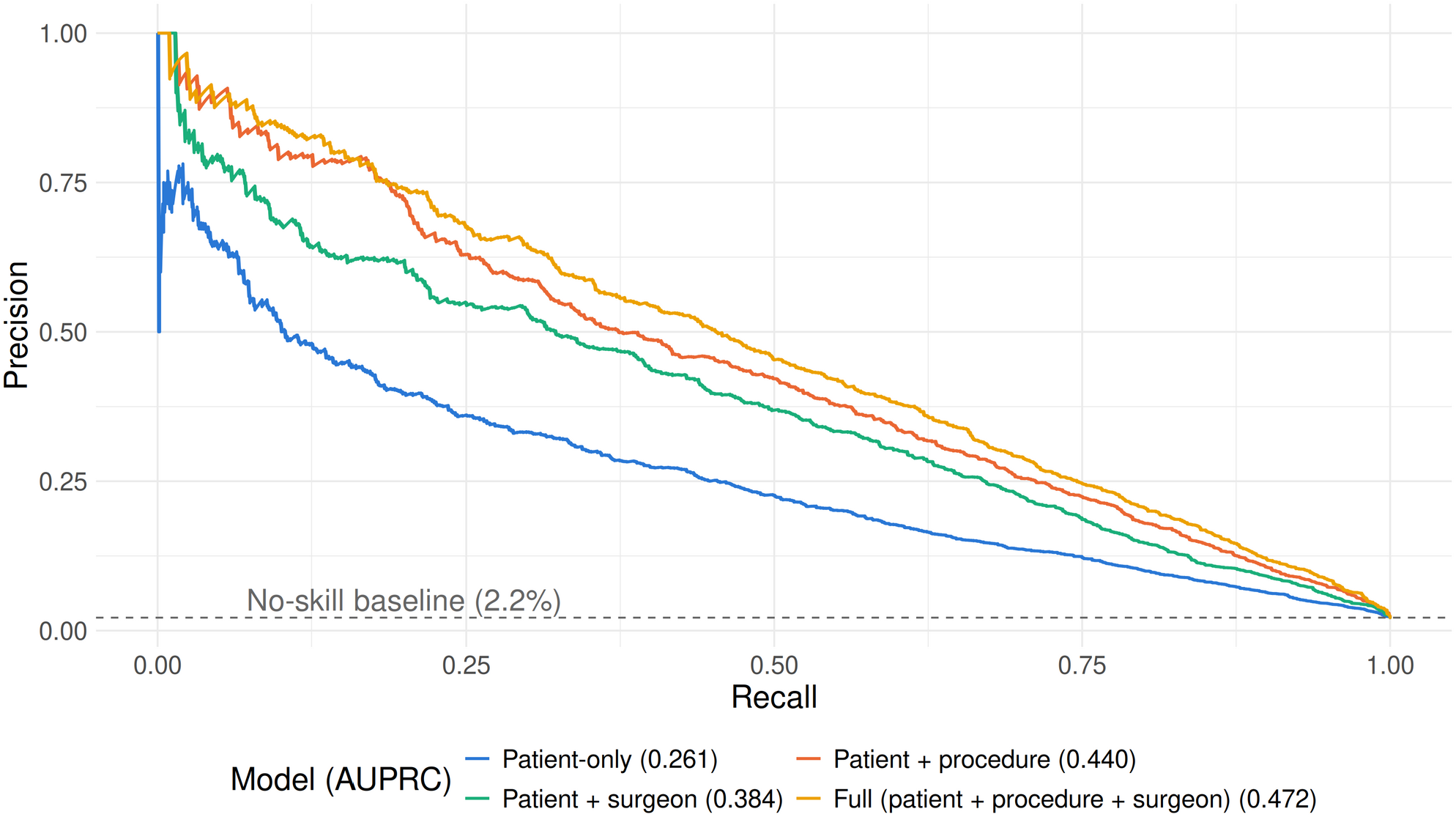
Precision-recall curves for the four CatBoost models on the held-out test set: the patient-only model (patient features only); the patient + procedure model (adding procedure group); the patient + surgeon model (adding surgeon identity); and the full model (patient + procedure + surgeon). AUPRC values are shown in parentheses in the legend. The dashed horizontal line indicates the no-skill baseline, equal to the test-set transfusion rate (2.2%). n = 55,980 procedures.

**Table 2.** Discrimination performance of the four CatBoost models on the held-out test set (n = 55,980 procedures). 95% CIs from a paired cluster bootstrap resampling patients (2,000 resamples). Lift = AUPRC divided by the no-skill baseline.

| Model | Grouping variables | AUPRC (95% CI) | Lift | ΔAUPRC vs patient-only (95% CI) | Brier score | AUROC (95% CI) |
|---|---|---|---|---|---|---|
| No-skill baseline | N/A | 0.022 | 1.0 | N/A | N/A | 0.5 |
| Patient-only | None | 0.261 (0.236–0.288) | 12.0 | 0 (reference) | 0.0182 | 0.90 (0.890–0.907) |
| Patient + procedure | Procedure group | 0.440 (0.409–0.471) | 20.3 | +0.178 (0.156–0.201) | 0.0155 | 0.94 (0.937–0.949) |
| Patient + surgeon | Surgeon | 0.384 (0.356–0.415) | 17.7 | +0.123 (0.103–0.144) | 0.0163 | 0.93 (0.924–0.938) |
| Full (patient + procedure + surgeon) | Surgeon + procedure | 0.472 (0.441–0.503) | 21.7 | +0.210 (0.185–0.235) | 0.0150 | 0.95 (0.944–0.955) |

### Model calibration

The full model achieved a Brier score of 0.015, calibration intercept 0.05 and slope 1.06, with predicted probabilities closely tracking observed event rates across deciles of the held-out test set (Supplementary Figure S3). Values for all four models are in Table 2 and Supplementary Table S4. Adding surgeon identity improved AUPRC in women and in all age groups, and less clearly in men (+0.014; 95% CI 0.000 to 0.029). Calibration was similar in all subroups (Supplementary Table S5).

### Permutation feature importance

In permutation feature importance for the full model (Figure 2), procedure group was the most important of the 72 features (mean AUPRC decrease 0.319 when shuffled), followed by preoperative haemoglobin (0.258) and surgeon identity (0.066; ranked third in 20/20 permutations). All other features had smaller effects (next largest: ASA class, 0.026). When surgeon identity was shuffled only within procedure groups, the AUPRC decrease was 0.062 (2.5th–97.5th percentile across permutations 0.055–0.069).

**Figure 2.**
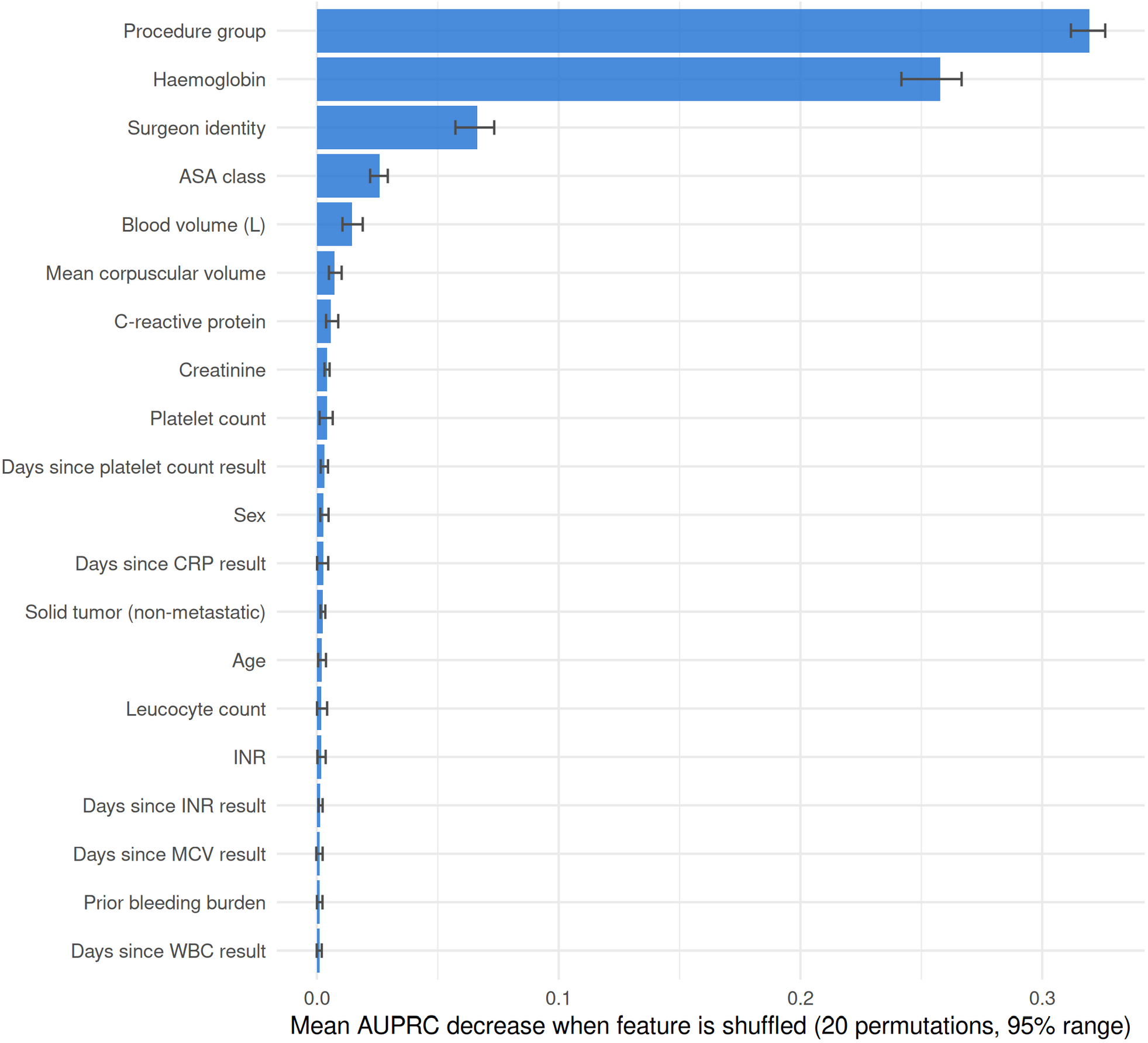
Permutation feature importance for the 20 most important features in the full model. Bars show the mean decrease in AUPRC when each feature’s values are randomly shuffled on the held-out test set, averaged over 20 permutations; error bars show the 2.5th–97.5th percentile range across permutations. Procedure group ranked first, preoperative haemoglobin second and surgeon identity third. All remaining features had mean decreases of 0.026 or less. n = 55,980 procedures.

### Surgeon-level variation

Transfusion rates varied substantially between surgeons within each of the ten most transfused procedure groups (Figure 3). Among the 749 surgeons with at least 15 test-set procedures, 96 had above-average and 14 below-average surgeon effects (mean SHAP contribution more than 1 SD above or below zero); the remaining 639 were within 1 SD (Figure 4). Procedures by above-average surgeons had 19% more transfusions than the patient + procedure model predicted (observed/expected 1.19; 95% CI 1.10–1.28; Supplementary Table S6). In exploratory analyses, the surgeon effect decreased with surgeon volume (Spearman ρ −0.72; 95% CI −0.77 to −0.67; Supplementary Figure S4) but differed little by seniority (Supplementary Methods, section 10).

**Figure 3.**
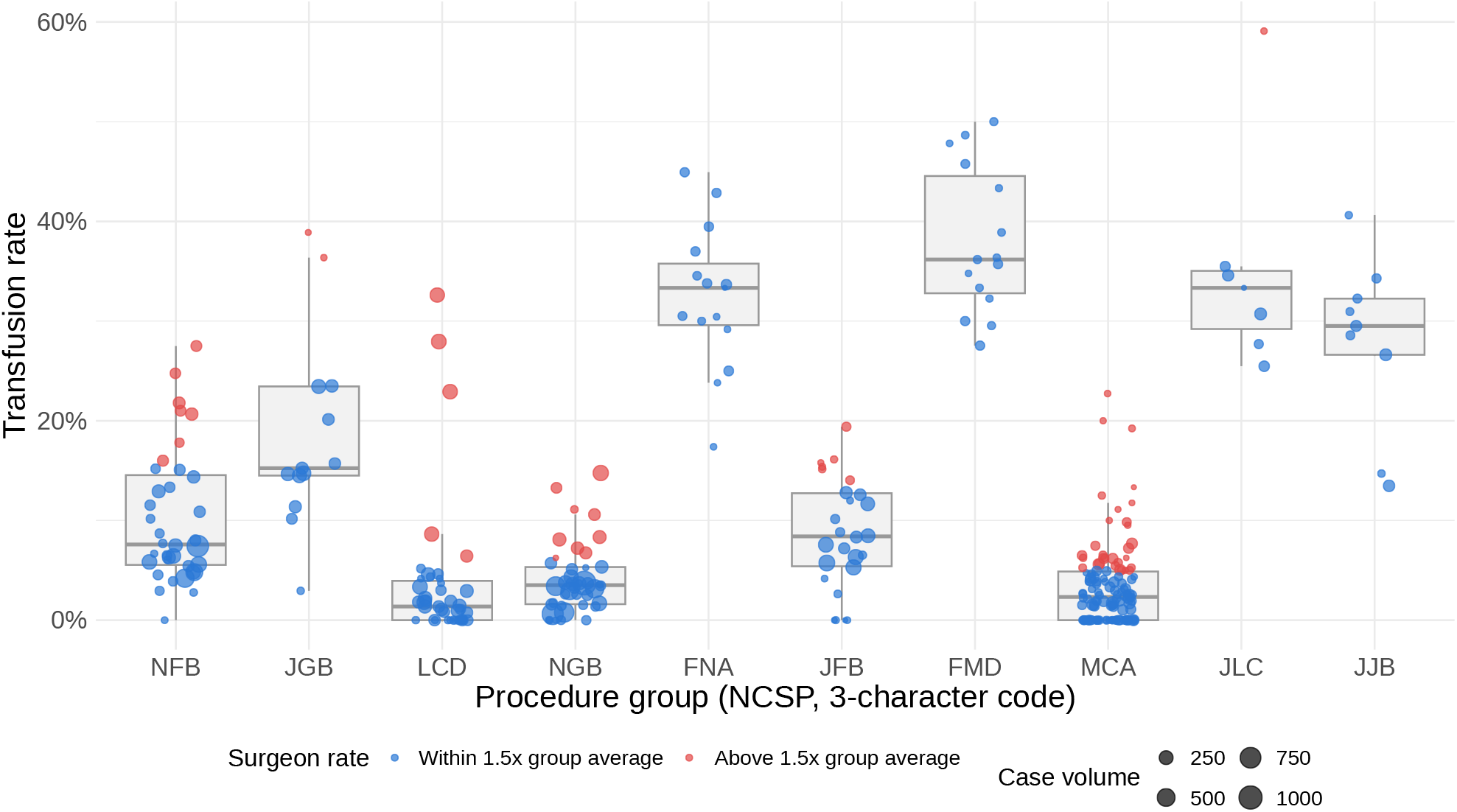
Per-surgeon transfusion rates within the ten most frequently transfused procedure groups. Each point represents one surgeon with at least 15 procedures in that group and point size is proportional to volume. Box-and-whisker plots show the distribution across surgeons (median, IQR, 1.5×IQR). Surgeons with a rate exceeding 1.5 times the group mean are highlighted in orange. Procedure groups are three-character NCSP designations (NFB: primary prosthetic replacement of hip joint, JGB: excision of rectum, LCD: total excision of uterus, NGB: primary prosthetic replacement of knee joint, FNA: connection to coronary artery from internal mammary artery, JFB: partial excision of intestine, FMD: replacement of aortic valve, MCA: caesarean section, JLC: excision of pancreas, JJB: resection of liver). n = 277 surgeons.

**Figure 4.**
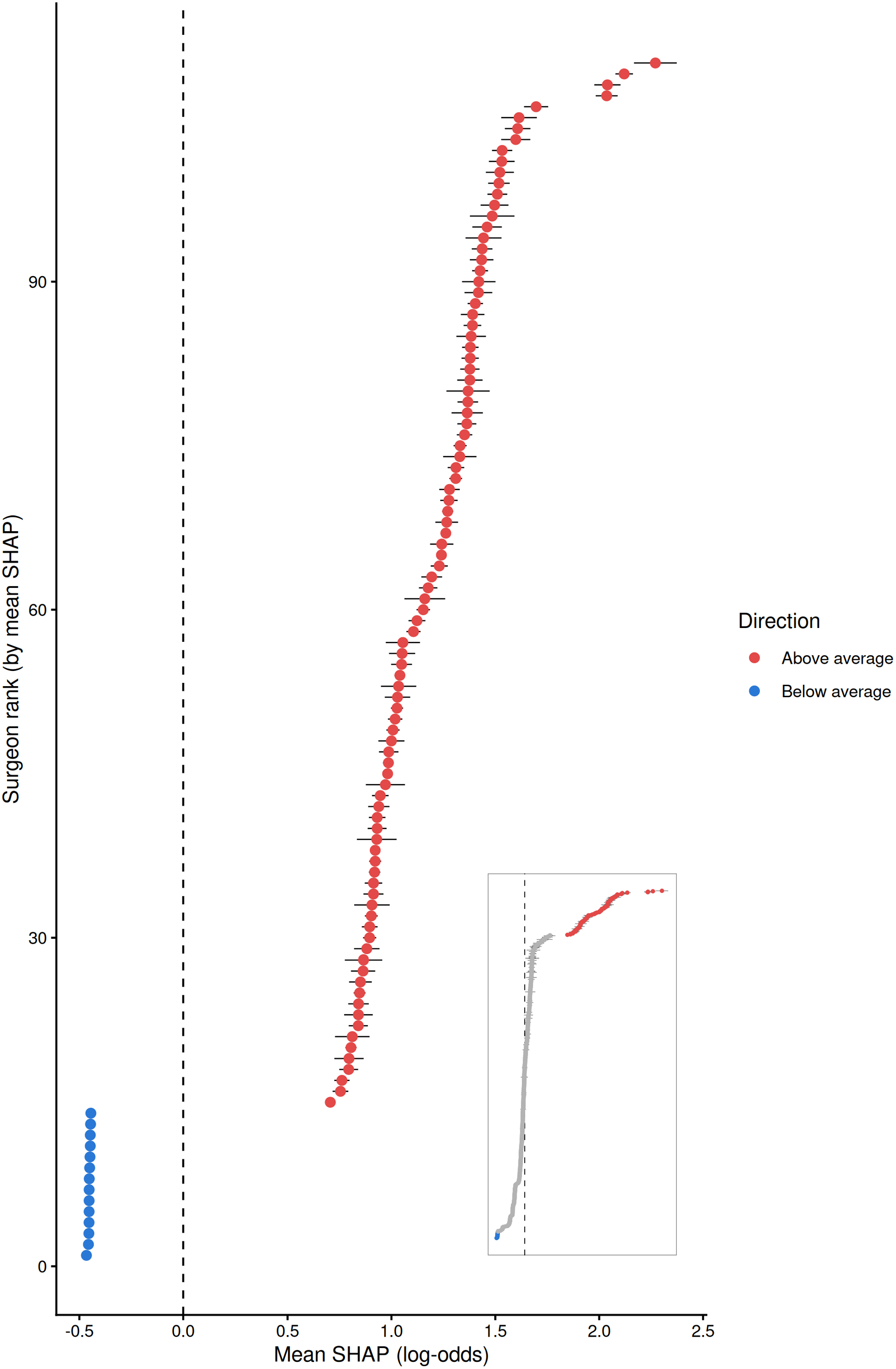
Surgeon-specific effects, estimated as each surgeon’s mean SHAP contribution for surgeon identity (log-odds scale) across their test-set procedures, among the 749 surgeons with at least 15 test-set procedures. The main panel shows the 110 surgeons whose mean SHAP contribution lay more than 1 SD above (red, n = 96) or below (blue, n = 14) zero. The inset shows all 749 surgeons, with the 639 surgeons within 1 SD in grey. Horizontal bars show ±1.96 SE of the surgeon’s SHAP contributions across their test procedures (consistency of the model’s attribution, not uncertainty in the surgeon effect). The dashed line at zero indicates no deviation from the average prediction.

## DISCUSSION

In this study, the operating surgeon was associated with RBC transfusion beyond patient factors and procedure type: adding surgeon identity to a model using data available seven days before elective surgery increased AUPRC by 0.032 (95% CI 0.020–0.044), raising lift over the no-skill baseline from 20.3 to 21.7. Surgeon identity was the third most important feature after procedure group and preoperative haemoglobin (Figure 2), consistent with a previous report.^11^ This variation plausibly reflects differences in practice, such as attention to haemostasis during surgery and thresholds for postoperative transfusion, although unmeasured case complexity may also contribute. The absolute gain is modest, but it came from a single routinely recorded variable. This matters especially for single-procedure transfusion risk models, which dominate the literature.

Systematic reviews classify transfusion prediction models simply as preoperative,^7^ but preoperative data can include results from the day of surgery.^11^ Correcting preoperative anaemia needs time for haemoglobin to respond to treatment,^38,39^ antithrombotic medication must be managed days in advance,^40,41^ and blood supply for elective lists is planned before the day of surgery. We therefore restricted all predictors to data recorded at least seven days before surgery, in line with PBM guidelines that emphasise early identification of high-risk patients.^42,43^ This has a cost: results from the final week, often including the most recent haemoglobin, were not available, so performance probably understates what a same-day model could achieve. Even so, preoperative haemoglobin was the strongest patient-level predictor, consistent with the central role of anaemia in PBM guidelines.^43^

The model also identified surgeons whose cases it consistently rated as higher or lower risk than those of colleagues performing the same procedures on similar patients, pointing to systematic differences in transfusion practice. These should not be read as differences in surgical skill, as some surgeons may take on more complex cases than the data can show. Recent consensus statements recommend routine audit of individual clinicians’ transfusion practice as part of PBM,^44^ and surgeon-level profiles could show where audit is most needed. They could also inform blood supply planning, since expected blood use for a scheduled elective list could take into account which surgeons are operating.^8^

In exploratory analyses, surgeons with higher procedure volume had lower surgeon effects, in line with studies linking surgeon experience and volume to fewer bleeding complications,^13,14,16,17^ whereas seniority made little difference. The decrease occurred in discrete steps likely because CatBoost’s default encoding includes how often each surgeon appears in the training data, so part of the surgeon effect is an implicit volume effect. Volume did not, however, account for the surgeons with clearly above-average effects (Supplementary Figure S4). Because surgeon volume and seniority are linked to case allocation within procedure groups, these associations cannot separate experience from case complexity. Their main value is to show whether the surgeon effect reduces to simple, measurable surgeon characteristics, or reflects individual practice that requires case-level audit.

We considered only the operating surgeon. Anaesthesiologist identity has previously been identified as a useful variable for type and screen order prediction^11^ and anaesthetic technique may also influence bleeding,^45,46^ although anaesthesiologists generally adhere to restrictive transfusion thresholds^47–49^ and most transfusions in surgical patients take place postoperatively on the wards.^50^ Future studies should include anaesthesiologist identity and surgeon–anaesthesiologist pairs, since complex cases may be assigned to particular pairs.

Although the cohort covers a regional network of hospitals, all data come from one health system in one country, with a shared EHR, transfusion policies and PBM culture, and the results may not generalise to settings with different case mixes, blood product availability or transfusion practices. Surgeon identity is also institution-specific, so other hospitals would need to retrain the model on their own data. Residual case-mix confounding cannot be excluded: unmeasured patient characteristics could partially explain the surgeon-level variation. Surgeon identity entered the model as a categorical predictor rather than as a level in a hierarchy of procedures, surgeons and hospitals, so it gives no uncertainty for individual surgeon effects; between-surgeon variation was estimated separately (Supplementary Table S7). Calibration varied between procedure groups, with caesarean section under-predicted (Supplementary Table S8). Finally, the model has not been externally validated, and prospective evaluation would be required before deployment.

In conclusion, the operating surgeon was associated with the likelihood of RBC transfusion in elective surgery, beyond what patient characteristics and procedure type explained. The association was detectable using only information available a week before surgery, and adding surgeon identity improved the prediction of transfusion from routinely collected data. Making surgeon-level differences visible at this point could support audit of transfusion practice as part of PBM and blood supply planning for elective surgical lists.

## ACKNOWLEDGMENTS

This study was supported by Government Research Funding (VTR) and the Finnish Red Cross Blood Service Research Fund as part of the Modelling and predicting blood product demand in HUS Helsinki University Hospital (HUS-RBC) research project. The funders had no role in the design of the study, the analysis and interpretation of the data, or the decision to publish.

## CRediT authorship contribution statement

**Esa Turkulainen:** Writing – Original Draft, Writing – Review & Editing, Software, Methodology, Investigation, Formal analysis, Data curation, Validation, Visualization, Conceptualization. **Mikko Arvas:** Writing – Review & Editing, Supervision, Funding Acquisition, Project Administration, Conceptualization. **Minna Ilmakunnas:** Writing – Review & Editing, Project Administration, Funding Acquisition. **Miika Koskinen:** Writing – Review & Editing, Project Administration, Methodology, Funding acquisition, Conceptualization.

## Data availability statement

The patient-level data cannot be shared publicly under the Finnish Act on the Secondary Use of Health and Social Data (552/2019). Researchers may apply for access to the data from the data controller, HUS Helsinki University Hospital. The analysis code is openly available.^37^

