## Supplementary material for "Prediction of elective surgical red blood cell transfusion reveals surgeon-level variation"

### Supplementary Methods

#### 1. Data sources, linkage and cleaning

##### DATA ACCESS AND LINKAGE

Data were extracted from the hospital EHR (Apotti), and for prescriptions also from the legacy system (Uranus), by HUS data lake personnel. The investigators received only a pseudonymised extract restricted to adult patients and had no access to the wider EHR population. The operation, laboratory, diagnosis, prescription, measurement, bleeding and transfusion tables were linked by a pseudonymised patient identifier assigned within the data lake. No linkage to external registries was performed.

##### SETTING

The procedures in the analysed cohort were performed in 33 operating units of the HUS hospital network. The unit was not recorded for 165 procedures (0.1%). The largest unit contributed 11.7% of procedures, and 14 units each contributed more than 3%.

##### STUDY POPULATION

Supplementary Figure S2 shows the selection of procedures. Elective status was defined by national urgency codes. Operations without a recorded urgency code (213,882 of 696,189 operations in the source table) were excluded, together with those coded as non-elective, because their elective status could not be established. The minimum-volume criterion applied to the full primary NCSP code, not to the three-character procedure group. All included patients were adults (no procedure with age at surgery below 18 years).

##### CLEANING OF SOURCE DATA

*Operations.* The source operation table holds one row per operation and assigned staff member. Its 719,737 rows were collapsed to 696,189 operations, keeping the staff member with the most senior professional title as the primary surgeon (ties broken at random with a fixed seed). Four records that duplicated another record's patient and operating-room entry time were removed.

*Laboratory results.* Free-text status entries were set to missing. Left-censored results ("<X") were set to X/2 and right-censored results (">X") to X.

*Transfusions.* RBC units were identified from the ISBT 128 product codes of administered units, supplemented by the product name where the code was missing. Records without a unit number, and duplicate records of the same unit, were removed. A small number of missing transfusion start times were imputed from the observed distribution of transfusion durations.

*Prior bleeding.* Patients with no recorded earlier surgical bleeding were assigned as having zero cumulative blood loss (230,064 procedures, 82.2%).

##### OUTCOME AND PREDICTOR ASSESSMENT

Outcomes and predictors were extracted automatically from routinely recorded EHR data, identically for all patients and without reference to each other, so no blinding was required. The only predictor requiring subjective judgement is ASA class, assigned by the anaesthesiologist in routine care.

##### CHOICE OF PREDICTORS

Candidate predictors comprised the patient factors used in previous transfusion prediction models and in patient blood management guidelines (haemoglobin and other laboratory values, body size, sex, age, renal function, comorbidity, antithrombotic and haematinic medication, prior bleeding), extended to all such EHR domains available at the predictive horizon. No data-driven pre-selection was performed: all 70 patient-level candidate features, plus surgeon identity and procedure group where applicable, were entered into the models.

### 2. Imputation of body mass index and blood volume

BMI in the source data is calculated from recorded height and weight. We used the latest available patient height and weight measurements to calculate any missing BMI values. For each procedure, the patient's most recent measurement recorded at least seven days before operating-room entry was used, and blood volume was estimated from it (Nadler formula for BMI ≤ 30 kg/m$^2$, Lemmens–Bernstein–Brodsky formula above 30 kg/m$^2$).

No qualifying measurement existed for 20,031 procedures (7.2%). For these, BMI and blood volume were imputed by linear regression on age, sex, ASA class and procedure group, fitted on the procedures with observed values ($R^2$ 0.14 for body mass index and 0.46 for blood volume). Predictions were clipped to the 1st–99th percentile of observed values. The imputation models did not use the outcome. They were fitted on the whole cohort before the training–test split, so the test set contributed to the imputation of predictor values (but not to any outcome information).

### 3. Derivation of INR from thromboplastin time percentage

Where no direct INR measurement was available but a thromboplastin time percentage (TT%) had been measured, INR was derived from TT% with a log–log linear regression:

$$\log(\mathrm{INR}) = \beta_0 + \beta_1 \cdot \log(\mathrm{TT\%})$$

The model was fitted by ordinary least squares on all paired TT% and INR results drawn at the same time in the laboratory data, restricted to INR < 10 to exclude extreme anticoagulation values. The fit uses laboratory values only and no outcome information. The fitted coefficients were $\beta_0$ = 2.573 and $\beta_1$ = −0.561 (n = 105,760 pairs; $R^2$ = 0.971; residual standard error 0.054 on the log scale; Supplementary Figure S1). Derived values were then used in the same way as direct measurements when selecting the most recent qualifying INR result. Of the 127,273 INR values in the analysed cohort, 100,199 (78.7%) were derived from TT%.

### 4. Encoding of categorical predictors

The four categorical predictors (surgeon identity, procedure group, sex and ASA class) were passed to CatBoost (version 1.2.10) as categorical variables, and the package's default encoding was used. Sex, with two levels, is one-hot encoded under the default settings. Each of the other categorical predictors is replaced during training by numeric summary statistics. Two statistics were active in the fitted full model (target and frequency). For surgeon identity, the frequency statistic is a measure of surgeon volume, and it is discretised into at most 15 bins, which is seen in the stepwise relationship between surgeon volume and the surgeon effect in Supplementary Figure S4. Surgeons with no training cases receive the prior value of the target statistic and the lowest frequency.

Under the default settings, CatBoost can also construct the same statistics for combinations of up to four features while building the trees. A combination joins categorical predictors with each other or with binary splits of numeric predictors. The fitted full model used several dozen such combinations of both kinds, including surgeon identity × procedure group. The full model can therefore represent surgeon differences that vary between procedure groups. Where surgeon identity was combined with numeric splits, those differences may also vary between patient subgroups. The model comparisons in the main text measure all the predictive information carried by surgeon identity, however it was encoded.

### 5. Sample size and data partitioning

The study size was determined by the available data: all eligible elective procedures in the five-year study period. The training set contained 223,925 procedures in 164,787 patients, with 4,918 transfused procedures (2.2%), or 68.3 events per candidate feature for the 72-feature full model. The held-out test set contained 55,980 procedures in 41,198 patients, with 1,215 transfused procedures (2.2%). This gave a 95% confidence interval width of about ±0.012 for the primary AUPRC difference.

The five patient-grouped tuning folds within the training set each contained 44,701–44,826 procedures (32,956–32,958 patients; 970–998 transfused procedures). The training and test sets did not differ in setting, eligibility criteria, outcome definition or predictors. Their characteristics are compared in Supplementary Table S9. Of the 1,543 surgeons, 1,516 had training-set procedures; 27 appeared only in the test set.

### 6. Hyperparameter tuning and final model settings

Hyperparameters were selected separately for each of the four models by a 30-draw random search. The search ranges were learning rate log-uniform in $10^{-2}$–$10^{-0.5}$, tree depth 4–10, and L2 leaf regularisation 1, 3, 5 or 10. Each draw was evaluated by five-fold patient-grouped cross-validation within the training set, optimising AUPRC. Each fold used early stopping (50 rounds without improvement, maximum 2,000 iterations). The final number of iterations was the mean best iteration across folds, and each model was then refitted on the full training set (Supplementary Table S10). All other settings were CatBoost 1.2.10 defaults:

| Setting | CatBoost parameter | Value (default) |
|---|---|---|
| Loss function | loss_function | Logloss |
| Tree growth | grow_policy | SymmetricTree (oblivious trees) |
| Boosting scheme | boosting_type | Plain |
| Borders for numeric features | border_count | 254 |
| Missing numeric values | nan_mode | Min (smaller than all observed values) |
| One-hot encoding | one_hot_max_size | Categorical features with ≤2 levels |
| Categorical statistics | simple_ctr, combinations_ctr | Borders (target), Counter (frequency) |
| Bins per categorical statistic | CTR border_count | 15 |
| Maximum features per combination | max_ctr_complexity | 4 (Supplementary Methods, section 4) |
| Row subsampling rate | subsample | 0.8 |
| Random strength | random_strength | 1 |
| Leaf estimation | leaf_estimation_method | Newton (10 iterations) |

### 7. Class weighting: ablation and rationale

Gradient-boosted tree training under a weighted logistic loss has a population-optimal decision function that is a strictly monotonic transform of the true outcome probability. Rank-based metrics (AUROC, AUPRC) are therefore theoretically invariant to weighting, but the absolute predicted-probability scale is not: it inflates toward the up-weighted class. Since the intended use of this model includes individualised risk estimation, an ablation was conducted on the full model to test whether inverse-frequency class weighting provided a ranking benefit large enough to justify the resulting miscalibration.

The ablation was run entirely within the training set, reusing its patient-grouped cross-validation folds: fold 1 served as the ablation holdout, and folds 2–5 as the ablation training data and its tuning folds, so no patient contributed to both. The ablation holdout contained 44,701 procedures (977 transfused). The full model was tuned and fitted twice, once with inverse-frequency class weighting and once without, using the same tuning procedure as the main models. Ranking was compared by an external, unweighted AUPRC computed directly from the holdout predictions, matching the primary evaluation used throughout the study. Calibration was compared by Brier score and by mean predicted probability versus observed event rate on the same holdout (Supplementary Table S3).

Unweighted training gave a slightly higher AUPRC and substantially better calibration. Class weighting was therefore not used in any of the four models reported in the main text.

### 8. Calibration assessment

For each model, calibration on the held-out test set was summarised by four measures: the Brier score (mean squared difference between predicted probability and observed outcome), the ratio of observed to expected events (O/E, observed event rate divided by mean predicted probability), and the calibration intercept and slope.

A perfectly calibrated model has O/E 1, intercept 0 and slope 1. A slope above 1 indicates predictions that are slightly too conservative (spread too narrowly around the mean). A slope below 1 indicates predictions that are too extreme, as typically results from overfitting. 95% confidence intervals were obtained with the same paired patient-cluster bootstrap as for AUPRC (2,000 resamples of test-set patients).

For the full model, the held-out test set was also divided into ten bins of equal size, ranked by predicted probability. Within each bin, the mean predicted probability was plotted against the observed event rate, with a binomial 95% confidence interval on the observed rate (Supplementary Figure S3).

All four models were well calibrated (Supplementary Table S4). Calibration intercepts were close to zero (0.05–0.07; 95% CIs including or bordering zero) and O/E ratios close to one (1.04–1.06). Calibration slopes were slightly above one (1.04–1.10), indicating marginally conservative risk estimates rather than overfitting. Calibration within individual procedure groups is described in Supplementary Methods, section 11.

### 9. Sociodemographic subgroups

Transfusion rates differed by sex and age in this cohort (Table 1), so the full model was evaluated separately in women and men and in three age groups (<50, 50–69, ≥70 years). For each subgroup we report AUPRC, lift (AUPRC divided by the subgroup's transfusion rate), the AUPRC gain over the patient + procedure model, and the calibration measures of Supplementary Methods, section 8. 95% CIs come from patient-cluster bootstrap resampling within the subgroup (2,000 resamples). No fairness-specific methods (such as reweighting or group-specific thresholds) were applied during model development.

AUPRC depends on outcome prevalence, which rose with age (1.3% below 50 years, 3.0% at 70 years or older). Subgroup AUPRCs are therefore not directly comparable, and the maximum attainable lift falls as prevalence rises.

Adding surgeon identity to the patient + procedure model improved AUPRC in women and in all three age groups (Supplementary Table S5). The gain ranged from +0.014 (95% CI 0.000–0.029) in men to +0.070 (0.028–0.114) below 50 years. Calibration was similar across subgroups: O/E ratios were 0.98–1.09 and calibration slopes 1.01–1.08. Lift was somewhat lower in women (20.1) than in men (24.2), and lower at 70 years or older (18.2) than in the younger groups (22.2–22.8). AUPRC itself was highest in the oldest group (0.539).

### 10. Surgeon variation beyond patient and procedure factors

The surgeon effects in the main text (Figure 4) are SHAP attributions of the full model and describe what the model learned from the training data. They were checked against test-set outcomes, which the model never saw, in three ways.

##### OBSERVED VERSUS EXPECTED TRANSFUSIONS

For each surgeon-effect category, observed test-set transfusions were compared with those expected from the patient + procedure model. 95% CIs are exact Poisson intervals, treating the expected count as fixed. Surgeons with above-average effects had 19% more transfusions than expected (O/E 1.19; 95% CI 1.10–1.28), and 29% more within the ten most transfused procedure groups (1.29; 1.17–1.43). Surgeons within ±1 SD had fewer than expected (0.86; 0.78–0.94). The 14 surgeons with below-average effects had no transfusions in 5,439 test-set procedures, against about 9 expected. Against the full model, the O/E ratio of above-average surgeons was 0.99 (0.92–1.07) (Supplementary Table S6). Across the 749 surgeons, the mean SHAP contribution correlated with the surgeon's own test-set O/E ratio (Spearman ρ 0.25; 95% CI 0.18–0.32, bootstrap over surgeons). Because many surgeons had no test-set transfusions, observed counts were increased by 0.5 before computing this ratio.

Surgeons with above-average effects operated on procedures of higher expected risk (median expected risk per procedure 11.9%, against 0.7% for surgeons within ±1 SD and 0.2% for those below average; ρ between mean SHAP and expected risk 0.48; 0.40–0.54). They operated in slightly fewer procedure groups (median 11 against 12; ρ −0.19; −0.26 to −0.12). The below-average group consisted of high-volume practices (median 1,876 procedures) in low-risk procedures.

##### SURGEON SENIORITY

Among the same 749 surgeons, each surgeon was classified by their professional title: senior (specialist or consultant; n = 330), not senior (resident or non-specialist; n = 279), or unknown (n = 140). Median (IQR) surgeon effects (mean SHAP contribution, log-odds) were -0.02 (−0.08 to 0.09) for senior surgeons, 0.03 (−0.04 to 0.12) for non-senior surgeons and 0.05 (-0.02 to 0.11) for surgeons with unknown title, so the effect differed little by seniority. The primary surgeon of each procedure was selected by seniority of title (main text, Data sources and study population), so seniority is also linked to how cases were attributed to surgeons.

##### RANDOM-INTERCEPT MODELS

Test-set transfusion was modelled by logistic regression with the logit of a model's predicted probability as an offset and random intercepts for procedure group and surgeon (lme4 2.0.6). The surgeon random intercept measures how much surgeons differ in transfusion given their patients' predicted risk and their procedures. It is summarised by its standard deviation and the median odds ratio (MOR, between two randomly chosen surgeons for comparable patients). Surgeon terms were tested by likelihood ratio against the model without them, halving the p-value for a variance on the boundary. 95% CIs are profile-likelihood intervals.

With the patient + procedure model as offset, the surgeon SD was 0.55 (95% CI 0.44–0.66; MOR 1.69; $p < 0.001$). Within the ten most transfused groups it was 0.46 (MOR 1.56; $p < 0.001$). With an additional surgeon × procedure group term, the surgeon SD was 0.53 (0.39–0.65) and the surgeon × procedure group SD 0.23 (0.00–0.48). The differences between surgeons were thus largely consistent across each surgeon's procedure groups. With the full model as offset, the surgeon SD fell to 0.30 (MOR 1.33; $p = 0.003$), a 71% reduction in variance. Within the ten most transfused groups it fell to 0.20 (0.00–0.41; $p = 0.17$) (Supplementary Table S7).

##### WITHIN-PROCEDURE DISCRIMINATION

Averaged over the 49 procedure groups with at least five test-set transfusions (1,128 of 1,215 transfusions), weighting each group by its number of transfusions, within-group AUPRC rose from 0.483 with the patient + procedure model to 0.503 with the full model (+0.020; 95% CI 0.006–0.034). This is about two-thirds of the pooled gain reported in the main text. The gain was +0.014 (-0.002–0.032) in the ten most transfused groups and +0.028 (0.003–0.053) in the other 39. It was not concentrated in small procedure groups: by tertile of the procedure group's training-set volume, the pooled gain was +0.028 (0.013–0.044), +0.045 (0.018–0.072) and -0.011 (−0.052 to 0.042) from smallest to largest. The largest tertile comprises seven low-risk groups with only 123 test-set transfusions.

Together, these analyses show that surgeons do differ in transfusion beyond measured patient characteristics and procedure groups, that these differences resurface in unseen outcomes, and that the full model captures most of them.

A surgeon effect raises or lowers the risk of all of a surgeon's procedures together, so it changes the ranking of procedures within a single group little. This is why AUPRC gains within individual procedure groups are small and imprecise (Supplementary Table S8), even where surgeon variation is clearly present. Surgeons with large effects operated on higher-risk procedures, and harder cases within the same procedure group (which the data cannot identify) may contribute to their effects (main text, Discussion).

### 11. Heterogeneity across procedure groups

Performance and calibration of the full model were also examined within the ten procedure groups with the most transfused procedures, which are the groups shown in the main text Figure 3 (Supplementary Table S8). Within-group AUPRC depends strongly on the group's transfusion rate, which ranged from 3.2% to 40.3%, so lift is the more comparable measure. Within-group lift ranged from 1.7 to 7.4, far below the pooled lift of 21.7, because most of the pooled discrimination comes from distinguishing procedure types.

Calibration (Supplementary Methods, section 8) varied between procedure groups, although good overall. With the full model as offset, the procedure-group random intercept had an SD of 0.70 log-odds (MOR 1.94) across all groups (Supplementary Table S7).

Two groups were under-predicted in both the training data (in-sample) and the test data: caesarean section (O/E 2.00 and 1.99, respectively) and primary knee replacement (O/E 1.53 and 1.67, respectively). Caesarean section also showed little discrimination even in the training data (in-sample AUPRC 0.24 at a 3.2% transfusion rate). Its patients were mostly ASA 1–2 (87%), and 46% had no qualifying haemoglobin result. Transfusion after caesarean section is driven mainly by postpartum haemorrhage arising during delivery, and obstetric risk factors (placental abnormalities, previous caesarean section, multiple pregnancy, pre-eclampsia) were not among the predictors. In several other groups, test-set O/E ratios exceeded those in training (e.g. excision of pancreas, 1.03 against 1.40). This partly reflects the selection of the ten groups by their number of transfused procedures, a count that includes the test set. Before predictions are used for procedure-specific blood supply planning or patient blood management, groups such as these would need procedure-specific recalibration.

### 12. Additional information

#### PROTOCOL, REGISTRATION AND PATIENT INVOLVEMENT

The study was not registered, and no protocol was prepared. Patients and outside parties were not involved in the design, conduct or reporting of this study.

#### INTENDED USE

The model is intended to be applied automatically to the EHR record of each scheduled elective procedure seven days before surgery. It supports patient blood management teams in identifying patients for preoperative optimisation, and transfusion services in planning blood supply for elective lists. Users do not enter data manually. Interpreting the predicted probabilities requires no statistical expertise. Treatments given after the predictive horizon, such as preoperative correction of anaemia, were not modelled: they are the interventions the prediction is meant to prompt. Because surgeon identity is specific to the institution, the fitted model cannot be transferred to other hospitals, but the code to retrain it is made available (main text, reference 37).

#### REPORTING GUIDELINE

We followed the TRIPOD+AI guidance in study reporting (Collins GS, Moons KGM, Dhiman P, et al. TRIPOD+AI statement: updated guidance for reporting clinical prediction models that use regression or machine learning methods. BMJ 2024;385:e078378). The completed checklist is provided at the end of this Supplement.

### Supplementary Tables

**Table S1.** Medication classes, ATC codes and look-back windows. A class was flagged if any prescription in it was active during the window before the predictive horizon.

| Drug class | ATC codes | Window |
|---|---|---|
| Anticoagulants | B01AA, B01AB, B01AE, B01AF, B01AX | 10 days |
| Antiplatelet agents | B01AC | 10 days |
| Dual antiplatelet therapy | Aspirin (B01AC06) and a P2Y12 inhibitor (B01AC04, B01AC22, B01AC24) both active | 10 days |
| Non-steroidal anti-inflammatory drugs | M01A, M01B | 10 days |
| Corticosteroids (systemic and topical) | H02A, D07 | 30 days |
| Coagulation factor concentrates | B02BD | 30 days |
| Systemic haemostatic agents | B02BX | 30 days |
| Oral iron | B03AA, B03AB | 90 days |
| Intravenous iron | B03AC | 90 days |
| Erythropoiesis-stimulating agents | B03XA | 90 days |
| Cytotoxic agents | L01 | 90 days |
| Immunosuppressants | L04 | 90 days |

**Table S2.** The 31 Elixhauser comorbidity categories (Quan et al. ICD-10 coding algorithm). Each category was encoded as the number of hospital visits on which it was recorded within the twelve-month look-back window. All 31 categories occurred in the cohort.

| Category | Category |
|---|---|
| Congestive heart failure | Peptic ulcer disease, excluding bleeding |
| Cardiac arrhythmias | AIDS/HIV |
| Valvular disease | Lymphoma |
| Pulmonary circulation disorders | Metastatic cancer |
| Peripheral vascular disorders | Solid tumour without metastasis |
| Hypertension, uncomplicated | Rheumatoid arthritis/collagen vascular diseases |
| Hypertension, complicated | Coagulopathy |
| Paralysis | Obesity |
| Other neurological disorders | Weight loss |
| Chronic pulmonary disease | Fluid and electrolyte disorders |
| Diabetes, uncomplicated | Blood loss anaemia |
| Diabetes, complicated | Deficiency anaemia |

| | |
|---|---|
| Hypothyroidism | Alcohol abuse |
| Renal failure | Drug abuse |
| Liver disease | Psychoses |
| | Depression |

**Table S3.** Class-weighting ablation for the full model, evaluated on an ablation holdout within the training set (cross-validation fold 1).

| Arm | Cross-validated AUPRC | Holdout AUPRC | Mean predicted probability | Observed event rate | Brier score |
|---|---|---|---|---|---|
| Weighted (inverse-frequency) | 0.427 | 0.444 | 0.156 | 0.022 | 0.069 |
| **Unweighted** | **0.446** | **0.458** | **0.021** | **0.022** | **0.015** |

**Table S4.** Calibration of the four models on the held-out test set (n = 55,980 procedures; 1,215 transfused). 95% CIs from a patient-cluster bootstrap (2,000 resamples). O/E, observed to expected events.

| Model | Brier score | O/E | Calibration intercept | Calibration slope |
|---|---|---|---|---|
| Patient-only | 0.0182 (0.0172–0.0192) | 1.06 (1.00–1.11) | 0.06 (0.00–0.12) | 1.10 (1.06–1.14) |
| Patient + procedure | 0.0155 (0.0147–0.0164) | 1.05 (1.00–1.10) | 0.06 (0.00 to 0.13) | 1.07 (1.03–1.10) |
| Patient + surgeon | 0.0163 (0.0155–0.0172) | 1.05 (1.00–1.10) | 0.07 (0.00–0.13) | 1.04 (1.01–1.08) |
| **Full** | **0.0150 (0.0142–0.0158)** | **1.04 (0.99–1.09)** | **0.05 (−0.01 to 0.12)** | **1.06 (1.02–1.09)** |

**Table S5.** Performance of the full model by sex and age group on the held-out test set, with the AUPRC gain over the patient + procedure model. 95% CIs from a patient-cluster bootstrap within each subgroup (2,000 resamples). Lift = AUPRC divided by the subgroup transfusion rate.

| | | Discrimination | | | Calibration | | | |
|---|---|---|---|---|---|---|---|---|
| **Subgroup** | **Transfused** | **AUPRC** | **Lift** | **ΔAUPRC** | **O/E** | **Intercept** | **Slope** | **Brier score** |
| **Sex** | | | | | | | | |
| Female n = 31,014 | 720 (2.3%) | 0.466 (0.424–0.504) | 20.1 (18.2–21.9) | +0.045 (0.028–0.063) | 1.04 (0.98–1.11) | 0.05 (−0.03 to 0.14) | 1.05 (1.01–1.10) | 0.0161 (0.0150–0.0173) |
| Male n = 24,966 | 495 (2.0%) | 0.480 (0.432–0.525) | 24.2 (21.7–26.9) | +0.014 (0.000 to 0.029) | 1.04 (0.96–1.11) | 0.05 (−0.05 to 0.15) | 1.06 (1.01–1.12) | 0.0136 (0.0125–0.0147) |
| **Age group** | | | | | | | | |
| <50 years n = 14,964 | 190 (1.3%) | 0.290 (0.225–0.363) | 22.8 (17.8–28.8) | +0.070 (0.028–0.114) | 1.09 (0.95–1.23) | 0.10 (−0.06 to 0.25) | 1.00 (0.93–1.08) | 0.0104 (0.0090–0.0119) |

| | | | | | | | | |
|---|---|---|---|---|---|---|---|---|
| 50–69 years n = 19,909 | 399 (2.0%) | 0.444 (0.391–0.494) | 22.2 (19.5–25.1) | +0.029 (0.008–0.049) | 0.98 (0.90–1.07) | −0.02 (−0.15 to 0.09) | 1.07 (1.01–1.14) | 0.0143 (0.0129–0.0158) |
| ≥70 years n = 21,107 | 626 (3.0%) | 0.539 (0.497–0.581) | 18.2 (16.6–19.7) | +0.025 (0.010–0.040) | 1.06 (1.00–1.13) | 0.09 (0.00 to 0.19) | 1.08 (1.02–1.14) | 0.0189 (0.0175–0.0204) |

**Table S6.** Observed and expected transfusions in the test set by surgeon-effect category (the categories of Figure 4: mean SHAP contribution for surgeon identity more than 1 SD above or below zero, among the 749 surgeons with at least 15 test-set procedures). Expected transfusions are the sum of predicted probabilities; 95% CIs are exact Poisson intervals. The expected risk per procedure is the median, across surgeons, of the mean predicted probability of the patient + procedure model.

| | | | | | | O/E vs patient + procedure model | | |
|---|---|---|---|---|---|---|---|---|
| Surgeon effect | Surgeons | Procedures | Transfused | Median volume | Expected risk | All groups | Ten groups | Full model |
| **Above average** | **96** | **4,694** | **691** | **219** | **11.9%** | **1.19 (1.10–1.28)** | **1.29 (1.17–1.43)** | **0.99 (0.92–1.07)** |
| Within ±1 SD | 639 | 42,946 | 434 | 253 | 0.7% | 0.86 (0.78–0.94) | 1.05 (0.92–1.21) | 1.10 (1.00–1.21) |
| Below average | 14 | 5,439 | 0 | 1,876 | 0.2% | 0.00 (0.00–0.40) | — | 0.00 (0.00–0.56) |

**Table S7.** Variation between surgeons and between procedure groups in test-set transfusion beyond the models' predictions. Random-intercept logistic models with the logit of the stated model's predicted probability as offset and random intercepts for procedure group and surgeon. SD on the log-odds scale with 95% profile-likelihood CI; MOR, median odds ratio; p, likelihood-ratio test of the surgeon term (halved for the boundary). —, profile interval did not converge.

| | Surgeon | | | Procedure group | |
|---|---|---|---|---|---|
| Offset (expected risk) | SD | MOR | p | SD | MOR |
| **All test-set procedures (n = 55,980)** | | | | | |
| Patient + procedure model | 0.55 (0.44–0.66) | 1.69 | <0.001 | 0.86 (0.68–1.08) | 2.27 |
| Full model | 0.30 (—) | 1.33 | 0.003 | 0.70 (—) | 1.94 |
| **Ten most transfused groups (n = 7,130)** | | | | | |
| Patient + procedure model | 0.46 (—) | 1.56 | <0.001 | 0.22 (—) | 1.23 |
| Full model | 0.20 (0.00–0.41) | 1.21 | 0.17 | 0.24 (0.10–0.46) | 1.26 |

**Table S8.** Performance and calibration of the full model within the ten procedure groups with the most transfused procedures (test set). 95% CIs from a patient-cluster bootstrap within each group (1,000 resamples). Lift = AUPRC divided by the group's transfusion rate. O/E in training is in-sample. The groups were selected by their number of transfused procedures in the whole cohort, which inflates test-set O/E ratios (Supplementary Methods, section 11).

| | | | | | O/E | |
|---|---|---|---|---|---|---|
| **Procedure group** | **Procedures** | **Transfused** | **AUPRC** | **Lift** | **Test** | **Training** |
| NFB<br>Primary prosthetic replacement of hip joint | 1,211 | 123 (10.2%) | 0.507<br>(0.418–0.596) | 5.0 | 1.19<br>(1.02–1.37) | 1.09 |
| JGB<br>Excision of rectum | 410 | 74 (18.0%) | 0.547<br>(0.433–0.662) | 3.0 | 1.03<br>(0.85–1.21) | 0.96 |
| LCD<br>Total excision of uterus | 1,178 | 79 (6.7%) | 0.499<br>(0.385–0.609) | 7.4 | 1.12<br>(0.92–1.34) | 1.01 |
| NGB<br>Primary prosthetic replacement of knee joint | 1,711 | 68 (4.0%) | 0.277<br>(0.171–0.410) | 7.0 | 1.67<br>(1.30–2.07) | 1.53 |
| FNA<br>Connection to coronary artery from internal mammary artery | 172 | 49 (28.5%) | 0.669<br>(0.534–0.783) | 2.3 | 0.89<br>(0.71–1.09) | 1.02 |
| JFB<br>Partial excision of intestine | 659 | 58 (8.8%) | 0.402<br>(0.272–0.530) | 4.6 | 0.91<br>(0.73–1.11) | 0.90 |
| FMD<br>Replacement of aortic valve | 135 | 51 (37.8%) | 0.787<br>(0.686–0.869) | 2.1 | 1.17<br>(0.95–1.36) | 1.06 |
| MCA<br>Caesarean section | 1,385 | 45 (3.2%) | 0.066<br>(0.034–0.138) | 2.0 | 1.99<br>(1.41–2.65) | 2.00 |
| JLC<br>Excision of pancreas | 129 | 52 (40.3%) | 0.700<br>(0.568–0.821) | 1.7 | 1.40<br>(1.14–1.65) | 1.03 |
| JJB<br>Resection of liver | 140 | 38 (27.1%) | 0.553<br>(0.404–0.708) | 2.0 | 1.14<br>(0.87–1.42) | 1.06 |

**Table S9.** Characteristics of the training and held-out test sets, including the share of procedures without a qualifying preoperative laboratory result. Values are n (%) or median (IQR).

| Variable | Training set | Test set |
|---|---|---|
| **Cohort** | | |
| Procedures | 223,925 | 55,980 |
| Patients | 164,787 | 41,198 |
| Transfused procedures | 4,918 (2.2%) | 1,215 (2.2%) |
| **Patient characteristics** | | |
| Age, years | 64 (48–75) | 64 (49–75) |
| Sex, female | 124,571 (55.6%) | 31,014 (55.4%) |
| Sex, male | 99,354 (44.4%) | 24,966 (44.6%) |
| ASA class 1 | 28,657 (12.8%) | 7,040 (12.6%) |

| | | |
|---|---|---|
| ASA class 2 | 101,467 (45.3%) | 25,264 (45.1%) |
| ASA class 3 | 67,741 (30.3%) | 17,178 (30.7%) |
| ASA class 4 | 9,130 (4.1%) | 2,215 (4.0%) |
| ASA class missing | 16,930 (7.6%) | 4,283 (7.7%) |
| Body mass index, kg/m² | 27 (24–30) | 27 (24–30) |
| Estimated blood volume, L | 4.8 (4.1–5.5) | 4.9 (4.1–5.5) |
| Haemoglobin, g/L | 136 (126–146) | 136 (126–146) |
| Platelet count, ×10⁹/L | 250 (208–298) | 249 (207–298) |
| Creatinine, µmol/L | 71 (61–85) | 71 (61–85) |
| van Walraven comorbidity index | 0 (0–4) | 0 (0–4) |
| Any recorded prior surgical bleeding | 40,081 (17.9%) | 9,760 (17.4%) |
| Anticoagulant | 25,879 (11.6%) | 6,322 (11.3%) |
| Antiplatelet agent | 14,869 (6.6%) | 3,733 (6.7%) |
| Oral or intravenous iron | 4,189 (1.9%) | 1,011 (1.8%) |
| **No qualifying preoperative result** | | |
| Haemoglobin | 69,474 (31.0%) | 17,673 (31.6%) |
| Platelet count | 69,486 (31.0%) | 17,681 (31.6%) |
| Leucocyte count | 69,475 (31.0%) | 17,673 (31.6%) |
| MCV | 69,476 (31.0%) | 17,673 (31.6%) |
| Creatinine | 76,743 (34.3%) | 19,396 (34.6%) |
| CRP | 113,976 (50.9%) | 28,843 (51.5%) |
| INR | 121,994 (54.5%) | 30,638 (54.7%) |
| Albumin | 189,455 (84.6%) | 47,611 (85.1%) |
| Ferritin | 201,914 (90.2%) | 50,670 (90.5%) |

**Table S10.** Final hyperparameters of the four models, selected by 30-draw random search with five-fold patient-grouped cross-validation on the training set (Supplementary Methods, section 6).

| Model | Learning rate | Tree depth | L2 leaf regularisation | Iterations | Cross-validated AUPRC, mean (SD) |
|---|---|---|---|---|---|
| Patient-only | 0.0159 | 9 | 3 | 1062 | 0.267 (0.018) |
| Patient + procedure | 0.0159 | 9 | 3 | 1033 | 0.425 (0.019) |
| Patient + surgeon | 0.0159 | 9 | 3 | 938 | 0.383 (0.020) |
| **Full** | **0.0159** | **9** | **3** | **809** | **0.457 (0.014)** |

### Supplementary Figures

**Figure S1.** Relationship between thromboplastin time percentage (TT%) and INR in paired same-draw measurements (log–log scale; INR < 10). The shaded density shows the paired measurements and the red line the fitted regression, log(INR) = 2.573 − 0.561 · log(TT%) (n = 105,760 pairs; $R^2$ = 0.971).

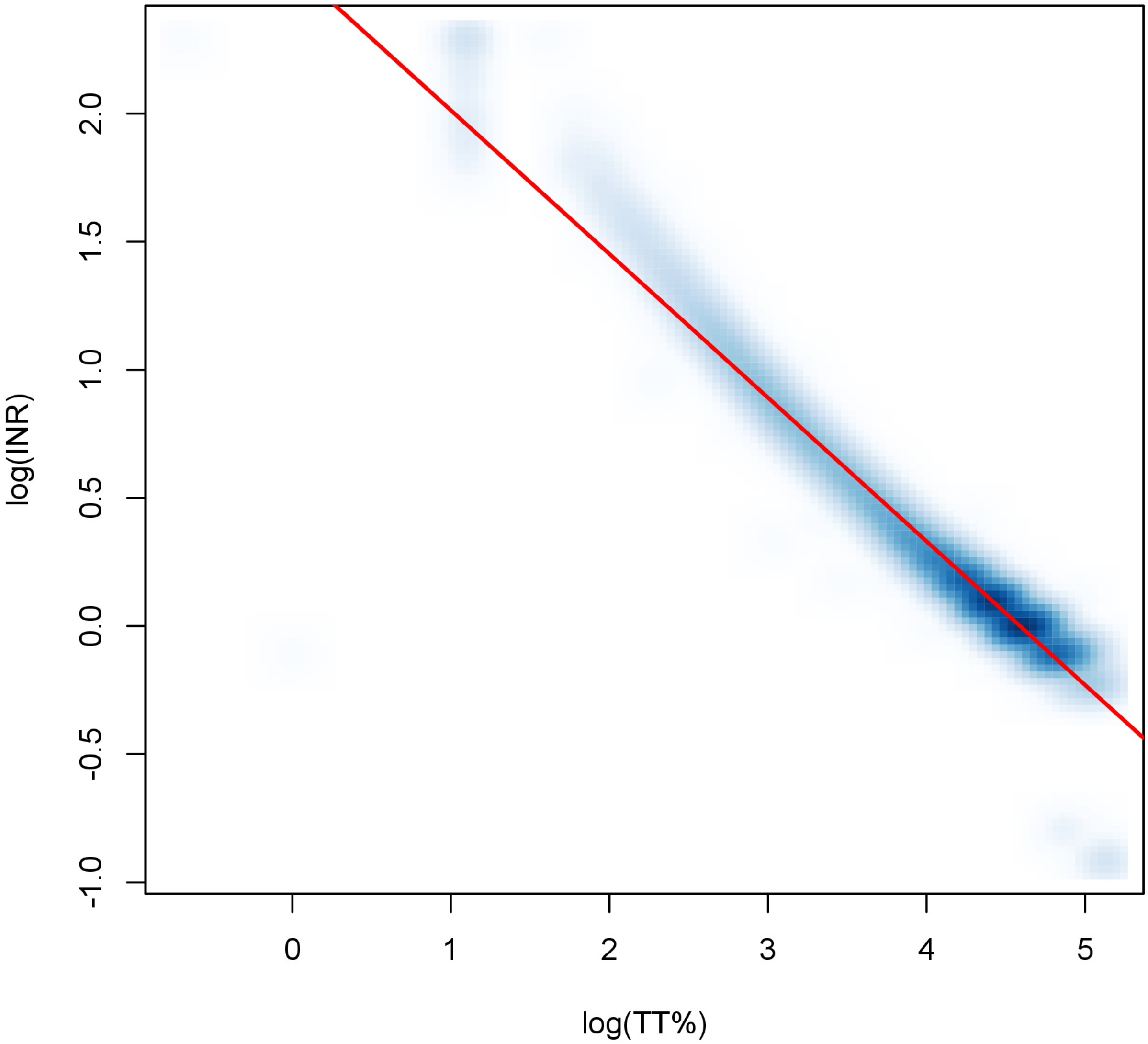

**Figure S2.** Selection of the study cohort and its partition into training and held-out test sets. Counts are procedures (patients). The five cross-validation folds used for hyperparameter tuning were formed from training-set patients only.

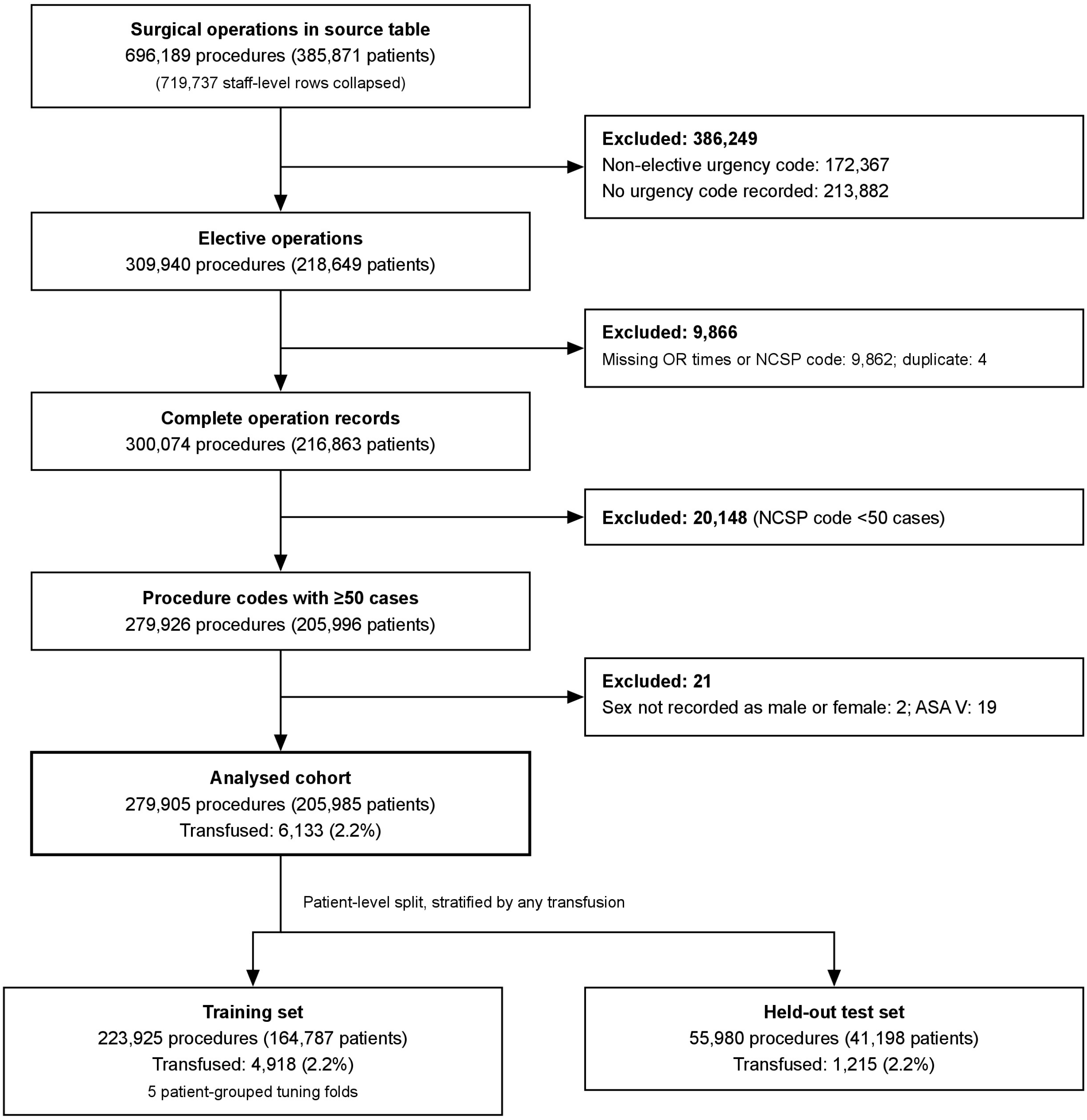

**Figure S3.** Calibration (reliability diagram) for the full model on the held-out test set. Points show mean predicted probability against observed event rate within each of ten rank-based bins of predicted probability, with vertical bars indicating binomial 95% confidence intervals on the observed rate. The dashed diagonal indicates perfect calibration. Brier score = 0.0150. n = 55,980 procedures.

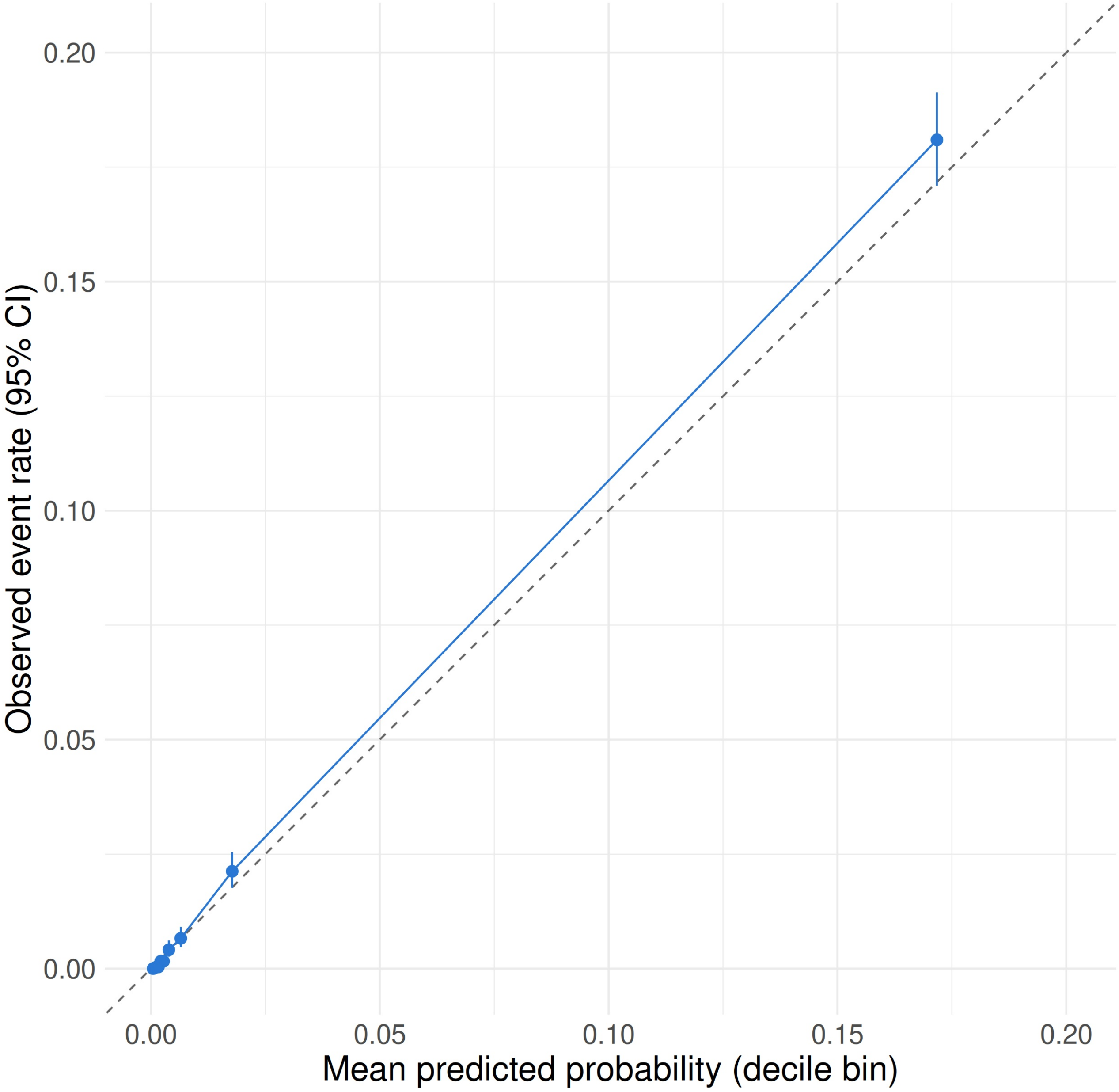

**Figure S4.** Surgeon effect versus surgeon volume (exploratory). Each point is one of the 749 surgeons with at least 15 test-set procedures. The x-axis shows the surgeon's total number of procedures in the study period (log scale), and the y-axis the surgeon's mean SHAP contribution for surgeon identity across their test-set procedures (log-odds scale). The red line is a smoother with 95% confidence band. Spearman ρ = −0.72 (bootstrap 95% CI −0.77 to −0.67). The stepwise pattern reflects CatBoost's default encoding of categorical predictors, which includes each surgeon's relative frequency in the training data, discretised into at most 15 bins.

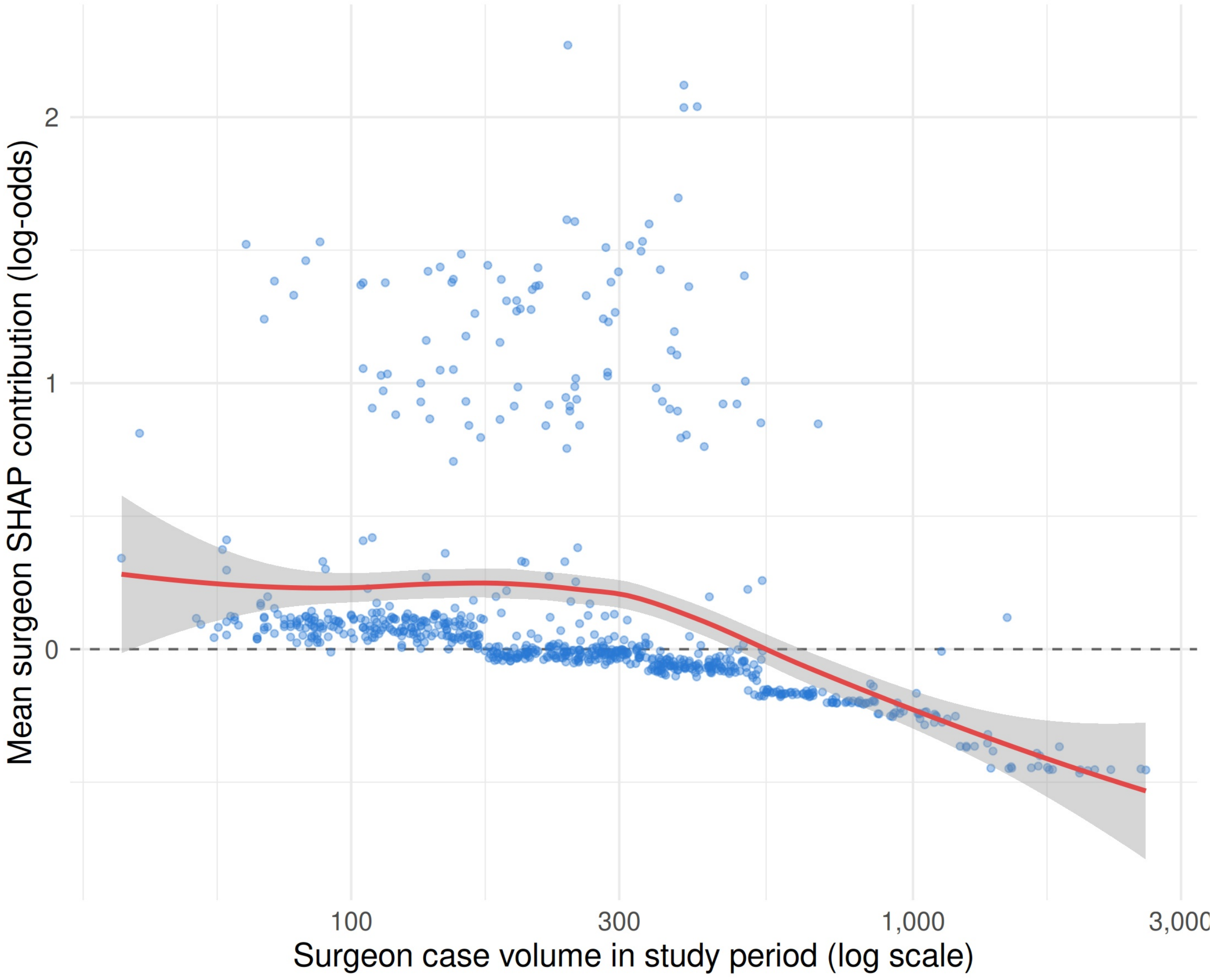
